# Association of HLA class I homozygosity with unfavorable clinical outcomes in patients with non-small cell lung cancer treated with PD-1/PD-L1 inhibitor as first-line therapy

**DOI:** 10.1101/2021.05.10.21256924

**Authors:** Dongyup Lee, Jonghanne Park, Horyun Choi, Gahyun Gim, Sukjoo Cho, Leeseul Kim, Youjin Oh, Cyra Y Kang, Yeseul Kim, Dean Tan, Pedro Antonio Hermida de Viveiros, Young Kwang Chae

## Abstract

**Background:** Homozygosity at HLA-I locus has been reported to be an unfavorable predictive biomarker of second-line or beyond immunotherapy in patients with different types of cancer. The linkage between HLA-I zygosity and survival in NSCLC patients treated with first-line immunotherapy with or without chemotherapy has not been reported.

**Methods:** Next generation sequencing with HLA genotyping was performed for patients with advanced NSCLC treated with immune checkpoint inhibitors with or without chemotherapy as first-line (N = 29). Progression free survival was compared between HLA-I homozygous (defined as homozygosity in at least one locus A, B, or C) and heterozygous patients. Kaplan-Meier curves were built, and log-rank test was used.

**Results:** Among 29 enrollees, 25 patients (86.2%) were HLA-I heterozygous and four patients (13.8%) were HLA-I homozygous. Treatment response was not available in five patients with HLA-I heterozygosity. Among 20 patients with HLA-I heterozygosity, five patients (20.0%) had partial response, 10 patients (50.0%) had stable disease, two patients (8.0%) had non-complete response/non-progressive disease, and three patients (12.0%) had progressive disease. Among four patients with HLA-I heterozygosity, one patient (25.0%) had partial response, one patient (25.0%) had stable disease, and two patients (50.0%) had progressive disease. The median progression free survival was not reached in heterozygous group and was 2.97 months in homozygous group (log-rank p = 0.68).

**Conclusions:** We observed a trend toward an inverse association between HLA-I homozygosity and survival outcomes in patients with NSCLC treated with first-line therapy containing immunotherapy. Further prospective studies to validate such a relationship are warranted.

## Introduction

With a better understanding of the immune evasion of cancer, immune check point inhibitors (ICIs) emerged to expand the therapeutic options for advanced cancer patients. By blocking cytotoxic T lymphocyte-associated protein 4 (CTLA-4) or the programmed cell death protein 1 (PD-1) or its ligand (PD-L1), ICIs counteract immune evasion of cancer cells through enhancing T cell mediated responses to cancer cell elimination ^1^.

On the basis of data providing survival benefit with an acceptable safety profile, ICIs became the new frontline treatment for advanced NSCLC ^2^. Pembrolizumab, an anti-PD-1 ICI, was approved by the U.S. FDA ^3^, as a single agent or in combination with chemotherapy, and as first-line treatment of patients with metastatic or unresectable NSCLC. However, only a subset of patients showed durable responses to ICIs, and treatment resistance is common ^4-6^. Thus, currently there exists a growing interest in establishing biomarkers that indicate treatment susceptibility and response.

To date, FDA-approved predictive biomarkers of response to ICIs are PD-L1 expression level and microsatellite instability (MSI) ^7,8^. In MSI-high tumors, a large number of somatic mutations are accumulated due to deficient DNA mismatch repair (MMR) gene ^9^. PD-L1 expression in tumor cells is positively correlated with response rates to anti-PD-1/PD-L1 therapy ^10^. Additionally, tumors with high TMB were shown to be more sensitive to ICI therapy ^11,12^. However, biomarkers in regard to the host germline genetics have not been investigated much.

Tumor-specific neoantigens have been shown to be of value in determining efficacy of tumor immune surveillance and predicting efficacy of ICIs ^13^. The human leukocyte antigen class I (HLA-I) plays a crucial role in presenting intra-cellular peptides on the cell surface for the recognition by T cell receptors. HLA downregulation has been found to be prevalent across a range of cancer types and has also been linked to poor outcome, possibly resulting reduced antigen presentation and thus facilitating immune evasion ^14,15^.

A recent large pan-cancer cohort study ^16^ investigated the association between HLA class I homozygosity and ICI treatment outcome. Homozygosity here was defined homozygosity in least one HLA-I locus (A, B, or C). The results demonstrated that HLA-I homozygosity was linked with shorter overall survival (OS). Furthermore, the study showed that the presence of HLA-I supertype B44 and absence of allele B15:01 were correlated with longer OS in a subgroup analysis of patients with melanoma. However, patients included in those studies were primarily those with melanoma. The correlation between HLA-I zygosity and tumor sensitivity to ICI therapy in patients with NSCLC still remains unclear ^9^.

Another study performed on patients with advanced NSCLC treated with ICIs showed no correlation between HLA-I zygosity and survival outcome ^17^. They investigated three cohort studies of 646 patients with advanced NSCLC who were treated with PD-1/PD-L1 or CTLA-4 inhibitor with or without chemotherapy. In the meantime, a meta-analysis of 19 randomized clinical trials involving 11,379 patients presented that anti-PD-1 provided better survival outcome when compared with that of anti-PD-L1 ^18^. Thus, we believed that it is meaningful to probe the correlation between HLA-I zygosity and survival outcome in a patient population with advanced NSCLC treated with frontline anti-PD-1 monotherapy with or without combination chemotherapy.

Herein, we investigated progression free survival (PFS) and OS in a cohort of patients with advanced NSCLC treated with ICIs with or without combination chemotherapy to delineate the correlation between HLA-I homozygosity and tumor sensitivity to ICI therapy. Also, we explored a subgroup of patients who were treated with chemoimmunotherapy to further clarify the correlation between HLA-I homozygosity and survival outcome.

## Materials and Methods

### Study oversight

A total of 29 patients treated with PD-L1 inhibitors underwent testing with Tempus (Tempus; Chicago, IL). Tempus is a Clinical Laboratory Improvement Amendments-certified and College of American-accredited method of sequencing both DNA and RNA that provides: genomic signatures, such as tumor mutational burden (TMB), and immune profiling including HLA genotypes and neo-antigen prediction. This method uses archival or fresh formalin-fixed paraffin embedded specimens isolated from patients’ tumor, blood or saliva specimens.

### Survival Analysis

PFS and OS were compared according to HLA-I homozygosity. PFS was defined as the time from the ICIs start date to cancer progression or death from any cause, and OS was defined as the from the ICIs start date to death from any cause. We investigated HLA-I variation at each locus (HLA-A, -B, and -C) and divided patients into two groups (HLA-I heterozygous in every loci vs. HLA-I homozygous in at least one locus). HLA genotypes were determined based on DNA sequencing results from patients’ blood. All patients underwent imaging studies before treatment and follow-up imaging studies 2 months after initiation of therapy. We calculated complete response (CR), partial response (PR), stable disease (SD), progressive disease (PD), non-CR/non-PD, and disease control rates (DCR) [CR, PR, and SD] for patients whose lesions are measurable according to response evaluation criteria in solid tumors (RECIST version 1.1) (N=24). Patients with CR, PR, non-CR/non-PD, or durable SD (SD after 6 months of treatment) were considered responder s; otherwise, they were considered non-responders. We also compared PFS differences according to TMB and PD-L1 expression level. High TMB was defined as TMB above the median. The expression of PD-L1 was stratified into three groups (<1%, 1%-49%, and ≥50%). PD-L1 immunostaining was performed in tumor tissues using SP142 antibody clones to evaluate PD-L1 expression level ^19^.

### Study participants and design

To study the association between HLA-I zygosity and survival outcome in patients, we retrospectively reviewed 80 patients with advanced NSCLC treated with ICIs with or without chemotherapy between September 2017 and March 2020. Only 29 patients with available next generation sequencing data (Tempus xT assay; Chicago, IL) were included. The HLA typing, neoantigen load, MSI status, and TMB were determined by DNA sequencing assay. HLA class I typing was acquired through Optitype in NGS data from RNA sequencing and exome sequencing 20,21. We also collected clinical data regarding survival outcome, in-house laboratory studies, and patient demographics. Patient demographic information was collected, including age, sex, diagnosis, performance status, BMI, smoking history, COPD, date of first and last ICIs treatment, date of progression, best overall response by RECIST (version 1.1) criteria, and if applicable, date of death. Key inclusion criteria was the availability of HLA genotype. Key exclusion criteria was any previous history of treatment for cancer being evaluated. Our study was performed in accordance with Northwestern University’s relevant guidelines/regulations and informed consent was obtained from all participants and/or their legal guardians. We received approval from Northwestern University Institutional Review Board and our study followed relevant guidelines and regulations.

### Statistical Analysis

Baseline characteristics of patient population are described below, and p-values displayed were calculated for the difference between the HLA-I homozygous group versus HLA-I heterozygous group. We performed Chi-square tests and Fisher’s exact tests for categorized variables and independent t-tests for continuous variables. P-values less than 0.05 were considered statistically significant. The patient survival according to HLA-I heterozygosity, TMB and PD-L1 were computed using Kaplan-Meier estimate with p-value determined by log-rank test. Using Cox proportional-hazards regression, we estimated the effect of HLA-I heterozygosity on survival outcome, adjusting for baseline patient characteristics including age, sex, and TMB to stratify randomization. Statistical analysis was performed using R software (version 1.2.5001).

## Results

### Patient population

We analyzed a total of 29 patients treated with first-line single-agent immunotherapy either with or without chemotherapy. The median duration of follow-up was 397 days (range 9 to 745 days). The median age was 72 years (age ranges from 55 to 90 years). Men were slightly more predominant (56.0%). Most patients (N = 22, 75.8%) had Zubrod performance status of 1-4. In terms of histology, 22 patients had non-squamous cell carcinoma (75.8%), and seven patients had squamous cell carcinoma (24.2%). Additionally, a majority of patients had stage 4 lung cancer (75.8%), while the remaining patients (24.2%) had stage 3 lung cancer. Three patients (10.3%) were never-smokers, and 26 (89.7%) were former or current smokers. The majority of patients were treated with pembrolizumab (N = 28, 96.6%), followed by atezolizumab (N = 1, 3.4%). Five of the patients (17.2%) were treated with ICIs monotherapy, while the remaining 24 patients (82.8%) were treated with combination chemoimmunotherapy.

Patients were divided into two groups based on HLA-I variation, HLA-I heterozygous (N = 25, 86.2%) and HLA-I homozygous (N = 4, 13.8%) (defined as homozygosity in at least one locus A, B or C). Baseline characteristics of patients were similar in two groups including age, gender, TMB and PD-L1. However, the number of neoantigens was statistically different (p = 0.006) between the groups. The mean number of neoantigens for HLA-I heterozygous patients were 6.5 compared to a mean number of neoantigens of 1.8 for HLA-I homozygous patients. For PD-L1 expression level, four (16.0%) HLA-heterozygous patients had PD-L1 level greater than or equal to 50%. In contrast, none of the patients from HLA-homozygous group had their PD-L1 level greater than or equal to 50%. Fourteen (56.0%) HLA-heterozygous patients had PD-L1 level that is greater than or equal to 1% but less than 50%, while two (50%) HLA-homozygous patients had their PD-L1 level in the same range. Seven (28.0%) patients from the HLA-heterozygous group and two (50%) patients from the HLA-homozygous group had PD-L1 level less than 1%. Four patients with HLA-I homozygosity received combination chemoimmunotherapy. Patient characteristics are described in Table A.

**Table A.** Characteristics of patient groups (N=29) with HLA-Heterozygous and HLA-Homozygous

|  | HLA-Heterozygous<br>(N=25) | HLA-Homozygous<br>(N=4) | P |
| --- | --- | --- | --- |
| ***General characteristic*** |  |  |  |
| Age, years | 69.7 ± 11.2 | 76.5 ± 6.2 | 0.25 |
| Male, No. (%) | 14 (56.0%) | 2 (50.0%) | 1.00 |
| BMI, kg/m<sup>2</sup> | 24.5 ± 5.2 | 23.1 ± 1.5 | 0.60 |
| Performance status |  |  | 0.50 |
| - 0, No. (%) | 5 (20.0%) | 2 (50.0%) |  |
| - 1-4, No. (%) | 20 (80.0%) | 2 (50.0%) |  |
| Smoking, pack-year (%) | 46.6 ± 40.0 | 31.2 ± 46.1 | 0.49 |
| COPD, No. (%) | 4 (16.0%) | 1 (25.0%) | 1.00 |
| Lung cancer type |  |  | 0.56 |
| - Non-squamous cell carcinoma, No. (%) | 18 (72.0%) | 4 (100.0%) |  |
| - Squamous cell carcinoma, No. (%) | 7 (28.0%) | 0 (0.0%) |  |
| Lung cancer stage |  |  | 1.00 |
| - Stage 1-3, No. (%) | 6 (24.0%) | 1 (25.0%) |  |
| - Stage 4, No. (%) | 19 (76.0%) | 3 (75.0%) |  |
| Best overall response |  |  | 0.37 |
| - Non-applicable, No. (%) | 5 (20.0%) | 0 (0.0%) |  |
| - Non-CR/Non-PD, No. (%) | 2 (8.0%) | 0 (0.0%) |  |
| - PD, No. (%) | 3 (12.0%) | 2 (50.0%) |  |
| - PR, No. (%) | 5 (20.0%) | 1 (25.0%) |  |
| - SD, No. (%) | 10 (40.0%) | 1 (25.0%) |  |
| ***Laboratory findings*** |  |  |  |
| Blood WBC count, x 10<sup>9</sup>/L | 9.1 ± 3.6 | 7.4 ± 2.7 | 0.39 |
| Blood Hemoglobin, g/dL | 11.8 ± 2.1 | 12.9 ± 0.8 | 0.32 |
| Blood Platelet count, x 10<sup>9</sup>/L | 345.3 ± 136.9 | 311.5 ± 39.0 | 0.63 |
| Blood Neutrophil count, % | 73.5 ± 13.0 | 70.1 ± 13.6 | 0.64 |
| Blood Lymphocyte count, % | 15.6 ± 10.2 | 18.5 ± 9.4 | 0.60 |

**Table A. continued**
|  | HLA-Heterozygous<br>(N=25) | HLA-Homozygous<br>(N=4) | P |
| --- | --- | --- | --- |
| ***Oncology testing panel findings*** |  |  |  |
| PD-L1 (IHC) |  |  | 0.55 |
| - <1%, No. (%) | 7 (28.0%) | 2 (50.0%) |  |
| - 1%<= <50%, No. (%) | 14 (56.0%) | 2 (50.0%) |  |
| - 50%<=, No. (%) | 4 (16.0%) | 0 (0.0%) |  |
| Tumor Mutation Burden, m/MB | 6.2 ± 4.4 | 2.3 ± 1.8 | 0.10 |
| Neoantigen, No. (%) | 6.5 ± 7.4 | 1.8 ± 1.3 | 0.01 |
| Frameshift mutations |  |  | 0.20 |
| - 0, No. (%) | 9 (36.0%) | 1 (25.0%) |  |
| - 1, No. (%) | 6 (24.0%) | 3 (75.0%) |  |
| - 2, No. (%) | 7 (28.0%) | 0 (0.0%) |  |
| - 3, No. (%) | 3 (12.0%) | 0 (0.0%) |  |

Abbreviations: BMI, body mass index; COPD, chronic obstructive pulmonary disease; PD, progressive of disease; PR, partial response; SD, stable disease; WBC, white blood cell; PD-L1, programmed death-ligand1; IHC, immunohistochemistry;

### Treatment response

Treatment response and disease progression were evaluated in accordance with RECIST criteria. Among 29 enrollees, 25 patients were heterozygous (86.2%) and four patients were homozygous (13.8%). Overall, five HLA-heterozygous patients were classified as non-applicable. In heterozygous group (N = 20), five patients (25.0%) had PR, 10 patients (50.0%) had SD, two patients (8.0%) had non-CR/non-PD, and three patients (15.0%) had PD. In homozygous group (N = 4), one patient (25%) had PR, one patient (25%) had SD, and two patients (50%) had PD. A total of five patients were observed to have PD, three HLA-heterozygous (15.0%) and two HLA-homozygous (50.0%). Six patients had PR, five HLA-heterozygous (20.0%) and one HLA-homozygous (25.0%). Eleven patients had SD, 10 HLA-heterozygous (40.0%) and one HLA-homozygous (25.0%). The DCR of patients treated with ICIs was 70%. In heterozygous group, the DCR was 72%, compared to 50% in homozygous group.

Also, 10 patients were classified as responders, while 14 patients were classified as non-responders. The median PFS of all 24 RECIST evaluable patients was 13.6 months (95% CI, 8.9 to could not be estimated), but the median OS was not reached (95% CI, 14.2 to could not be estimated). No statistically significant differences in OS and PFS were observed between responders and non-responders (log-rank p = 0.10, both) (see Figure, Supplemental Data E.1 and E.2), but showed trends for better survival in responder group. Among responders, one was treated with ICIs monotherapy, and nine were treated with combination chemoimmunotherapy. Three non-responders were treated with ICIs monotherapy, and 11 with combination chemoimmunotherapy. For PD-L1 expression status, one (10.0%) responder had PD-L1 level greater or equal to 50%, as opposed to 2 (14.3%) non-responders who had PD-L1 level greater or equal to 50%. The PD-L1 level of four (40.0%) responders ranged between 1% and 50%, while that of eight (57.1%) non-responders fell within the same range. The PD-L1 level of five (50.0%) responders was less than 1%, and that of four (28.6%) non-responders was less than 1.

### Survival analysis in patients treated with first-line immunotherapy with or without chemotherapy

The median PFS in the HLA-I heterozygous patient group was prolonged when compared with the HLA-I homozygous patient group (log-rank p = 0.68, Figure A). Patients with HLA-I homozygosity demonstrated shorter PFS (HR = 1.4, 95% CI = 0.30-6.4, p = 0.68). There was no significant difference between the two groups in terms of OS (log-rank p = 0.14). The HR in the analysis of OS was incalculable. There seemed to be a trend for shorter PFS in homozygosity group. However, both PFS and OS analysis showed no statistically significant correlation with the PD-L1 level or the TMB level. No statistically significant difference was noted between groups stratified by the different PD-L1 expression levels (<1%, 1%-49%, and ≥50%) (PFS log-rank p=0.65, OS Log-rank p = 0.10) (see Figure, Supplemental Data A.1 and A.2). Similarly, the difference between a group with the TMB level above the median and a group with the TMB level below the median was not statistically significant. (PFS log-rank p = 0.49, OS Log-rank p = 0.31) (see Figure, Supplemental Data C.1 and C.2). In a multivariable analysis adjusting for age (adjusted HR = 1.5, 95% CI = 0.30-7.3, p = 0.63), sex (adjusted HR = 1.3, 95% CI = 0.29-6.3, p = 0.70), and TMB (adjusted HR = 1.1, 95% CI = 0.21-5.6, p = 0.92), there was no statistical significance.

**Figure A.**
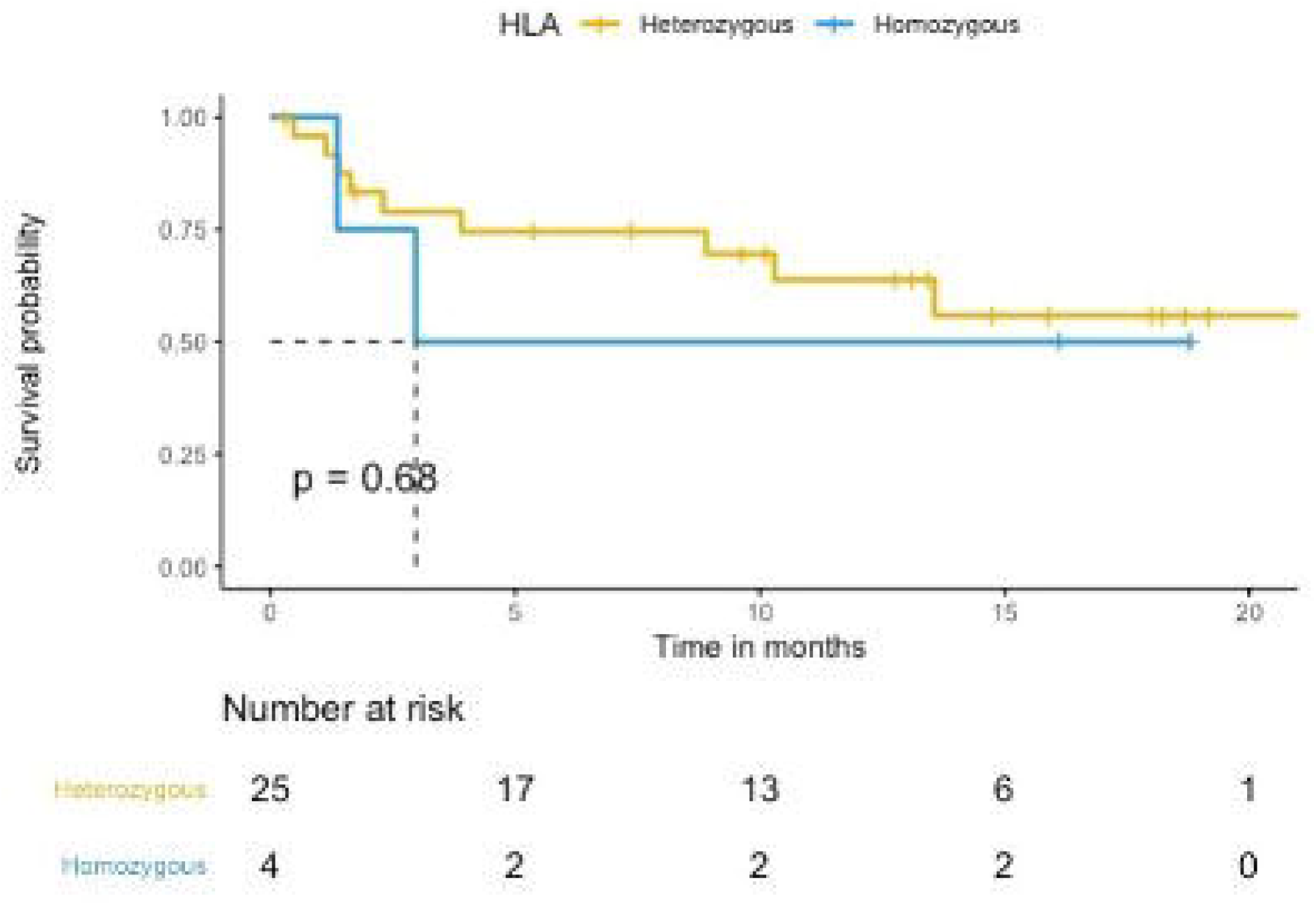
PFS according to HLA zygosity (N=29). Patients treated with first-line immunotherapy with or without chemotherapy (N=29). The median PFS in the HLA-I heterozygous patient group was prolonged when compared with the HLA-I homozygous patient group.

### Survival analysis in patients treated with first-line combination chemoimmunotherapy

The median PFS in the HLA-I heterozygous patient group was prolonged when compared with the HLA-I homozygous patient group (log-rank p = 0.79, Figure B). Patients with HLA-I homozygosity demonstrated shorter PFS (HR = 2.1, 95% CI = 0.55-8.2, p=0.27). There was no significant difference between the two groups in terms of OS (log-rank p = 0.19). The HR in the analysis of OS was incalculable. There seems to be a trend for shorter PFS in homozygosity group. However, both PFS and OS analysis showed no statistically significant correlation. No statistically significant difference was noted in PFS and OS among groups with different PD-L1 levels (PD-L1 was divided into three groups; <1%, 1%-49%, and ≥50%) (PFS log-rank p = 0.62, OS log-rank p = 0.21) (see Figure, Supplemental Data B.1 and B.2) and TMB groups (TMB above the median vs below the median) (PFS log-rank p = 0.80, OS log-rank p = 0.68) (see Figure, Supplemental Data D.1 and D.2). In a multivariable analysis adjusting for age (adjusted HR = 2.4, 95% CI = 0.59-10, p = 0.22), sex (adjusted HR = 2.1, 95% CI = 0.55-8.1, p = 0.28), and TMB (adjusted HR = 1.4, 95% CI = 0.33-5.9, p = 0.66), there was no statistical significance.

**Figure B.**
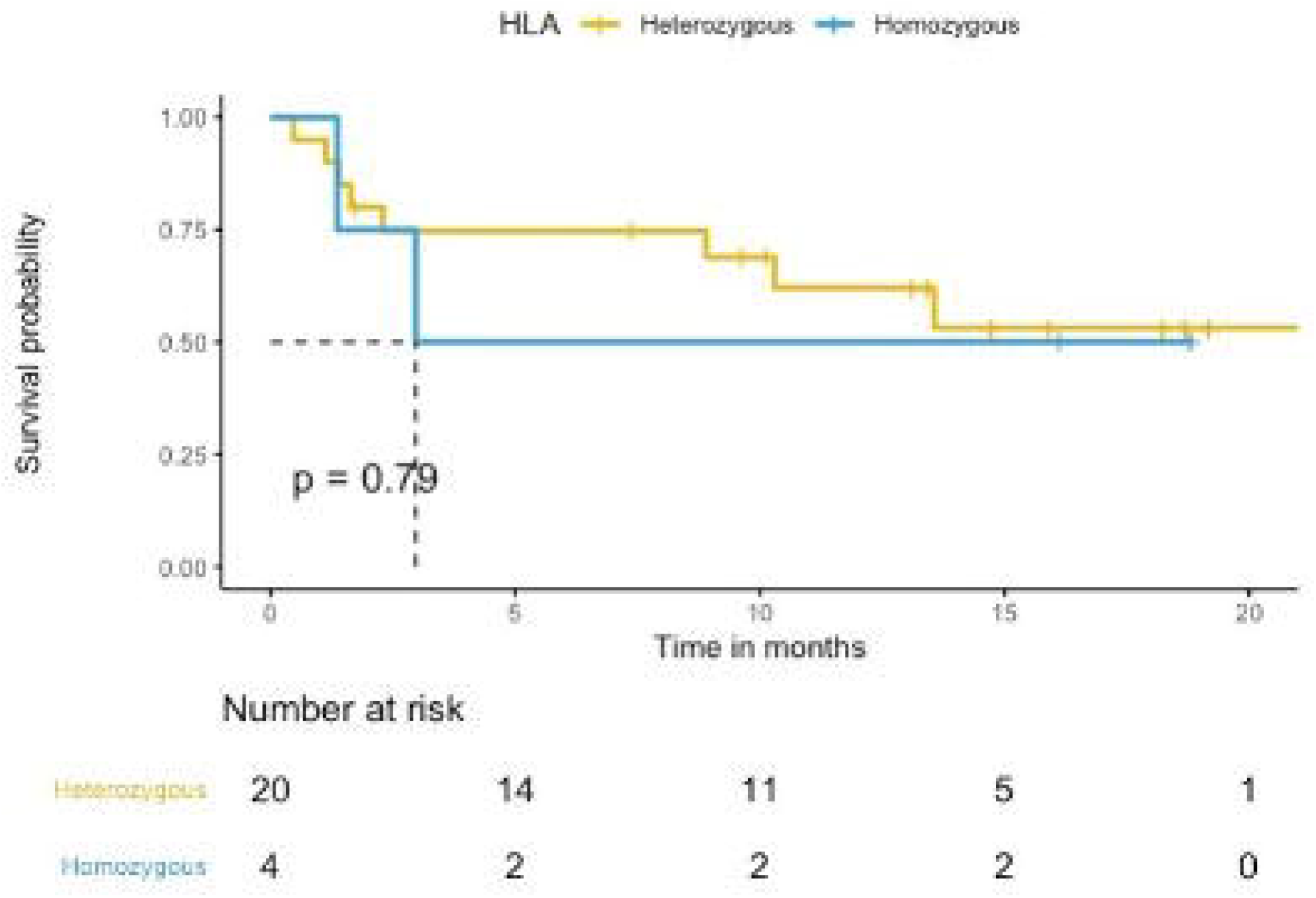
PFS according to HLA zygosity (N=24). Patients treated with first-line combination chemoimmunotherapy (N=24). The median PFS in the HLA-I heterozygous patient group was prolonged when compared with the HLA-I homozygous patient group

### HLA-I supertypes

Prior studies have shown that HLA-I alleles can be classified into 12 supertypes according to similar peptide binding affinities [15]. Therefore, we grouped patients’ HLA class I A and B alleles into 12 supertypes (supertype A01, A02, A03, A01-A03, A01-A24, A24, B07, B08, B27, B44, B58, and B62) and evaluated whether patients with any particular supertypes showed improved survival. In univariable analysis, none of the supertypes were correlated with PFS or OS. Patients HLA-I genotypes and supertypes were described (see Table, Supplemental Data B and C). Also, the PFS hazard ratios according to different HLA-I zygosities along with 95% confidence intervals were presented (see Table, Supplemental Data D).

## Discussion

Immunotherapy has drastically changed the treatment paradigm for patients with NSCLC. However, only some patients achieve durable survival outcomes. In order to ideally guide clinical practice, it is important to establish more accurate predictive biomarkers. Studies have shown that the antitumor activity of ICIs depends on HLA class I–related immune activity, and resistance to immunotherapy is associated with defective antigen presentation mediated by HLA class I [15 16]. Therefore, the HLA class I zygosity can serve a potential predictive biomarker for immunotherapy response.

One study involving 1535 patients in the advanced stage of multiple types treated with ICIs demonstrated the association of HLA-I homozygosity with poor survival outcome [15]. Two sets of cancer patients were evaluated in this study. Cohort 1 involved 369 patients, including 269 patients with advanced melanoma and 100 patients with NSCLC, treated with anti-CTLA-4 or anti-PD-1. Cohort 2 involved 1,166 patients with diverse cancer types, including melanoma and NSCLC, treated with anti-PD-1/PD-L1, anti-CTLA-4, or a combination of both. HLA-I homozygosity and low TMB were associated with decreased OS in cohort 1 and 2 (HR = 2.03 and 2.98, respectively). Also, in two independent melanoma cohorts, HLA-B44 supertype were associated with increased survival and HLA-B62 supertype with decreased survival.

Another study involving patients with advanced NSCLC treated with ICIs with or without chemotherapy evaluated HLA-I zygosity as a prognostic biomarker for NSLCL [16]. Three cohort studies were evaluated in this study. First, in M. D. Anderson Cancer Center (MDACC) cohort, 200 NSLCL patients’ PFS and OS were evaluated, who were treated with PD-1/PD-L1 inhibitor with or without chemotherapy. Second, in the CheckMate 012 trial (CM 012) cohort, the safety and activity of combination nivolumab plus ipilimumab were evaluated in chemotherapy-naïve 75 NSCLC patients using PFS data. Nivolumab, a PD-1 antibody, and ipilimumab, a CTLA-4 antibody, work synergistically together to induce stronger anti-tumor response [21]. Lastly, in Chowell cohort, patients’ OS was evaluated, including 100 NSCLC patients who were treated with anti-CTLA-4 or anti-PD-1 therapy (mainly with anti-PD-1 monotherapy) and 271 NSCLC patients who were treated with anti-CTLA-4, PD-1/PD-L1 inhibitor, or combination. There was no significant correlation between HLA-I zygosity and survival outcome in three independent cohorts (HR = 0.87 in MDACC cohort, 0.86 in CM012 cohort, and 1.31 in Chowell cohort).

Notable differences existed between patient populations in previous studies and ours. MDACC cohort included patients who either received treatment before or whose history of treatment was not disclosed in CM 012 and Chowell cohorts. Also, in CM 012 cohort, patients were treated with combination nivolumab plus ipilimumab. Thus, it is difficult to measure the predictive value of HLA-I zygosity as a biomarker for response to PD-L1 inhibitor.

The validity of biomarker might be different between treatment naïve patient group and treatment experienced group. For this reason, we restricted our patient population to only include patients who were treated with frontline pembrolizumab or atezolizumab, which are both FDA-approved regimens [2 7]. Consequently, we tried to focus on real-world patient data which include standard of treatment. We noticed the correlation between HLA-I homozygosity and shorter PFS, but it was not statistically significant. Additionally, HLA-I homozygosity in at least one locus showed no correlation with prolonged survival, unlike previous pan-cancer study. The reason for this discrepancy is unclear, but heterogenous response rates to ICIs according to different cancer types may account for some of these differences. For example, it is known that the response rate to ICIs in patients with melanoma is higher. Based on the fact that anticancer activity of ICIs depends on CD 8 T-cell and antigen presentation to CD 8 T-cell is mediated by HLA-I molecule, the impact of HLA-I zygosity in patient survival might be heterogenous according to cancer types [22 23].

Patients who received combination chemotherapy and immunotherapy were only eight percent in MDMCC cohort, and none were treated with combination chemoimmunotherapy in CM 012 and Chowell cohort. Indeed, the impact of predictive biomarkers appears to be different in predicting survival benefit from ICIs when immunotherapy was combined with chemotherapy in patients with NSCLC. For example, trials of chemoimmunotherapy have shown beneficial effects regardless of PD-L1 expression level, with a greater benefit in patients with PD-L1 ≥50% with no regards to specific drugs used (either immunotherapy or chemotherapy) or histology (either squamous or non-squamous) [24-26]. Thus, we also focused on subgroup analysis in patients with combination chemoimmunotherapy and found no statistical significance. Subgroup characteristics are described in supplemental Table A.

There existed correlation between HLA-I homozygosity and shorter PFS, but it did not reach statistical significance in patients with advanced NSCLC. Also, HLA-I homozygosity in at least one locus showed no correlation with OS. One possible explanation for these results is that chemotherapy augments tumor immunity by altering immune function. Based on preclinical evidence, we understand that the combination between ICIs and chemotherapy may achieve additive or synergistic effect owing to immunological effect of cytotoxic agents through inducing PD-L1 expression on the surface of tumor cells, selective elimination of myeloid-derived suppressor cells, and promoting T-cell priming and recruitment to the cancer cells [27-29]. The result also may be attributable to limited sample size. In the meantime, however, subgroup analysis in patients who were treated with anti-PD-1 monotherapy was not possible due to the small sample size. Therefore, further large prospective studies with different combinations of regimen are warranted to establish ideal biomarkers for selection of candidates for ICI treatment.

A previous pan-cancer study classified melanoma patients’ HLA-I alleles into six A supertypes and B supertypes. It demonstrated the association of supertype B44 with increased survival outcome, and supertype B62 with decreased survival outcome [15]. However, we were limited by the sample size for such analysis. This requires further validation with a larger cohort study, especially in patients with NSCLC.

## Conclusion

We observed trends toward an inverse correlation between HLA-I homozygosity and survival outcomes in patients with NSCLC treated with frontline immunotherapy with or without chemotherapy. Due to limited sample size, our analysis failed to reach statistical significance. Further studies are imperative to clarify the role of HLA-I zygosity in immunotherapy response and how it differs by different cancer types and treatment regimens.

## Data Availability

We attached supporting data and/or materials with our publication

## Acknowledgements

We thank all participants and their families for participating in this study

## Author contribution

DL: Writing - Original Draft, Formal analysis, Investigation, JP: Supervision, Methodology, Writing - Review & Editing, Validation, HC: Formal analysis, Writing - Review & Editing, Investigation, GG: Writing - Review & Editing, Investigation, LK: Writing - Original Draft, YO: Investigation, Writing - Review & Editing, SC: Writing - Review & Editing, CYK: Investigation, Writing - Review & Editing, YK: Investigation, Writing - Review & Editing, DT: Review & Editing, PV: Writing - Review & Editing, YKC: Supervision, Conceptualization, Validation

## Funding

This research did not receive any specific grant from funding agencies in the public, commercial, or not-for-profit sectors.

We attached supporting data and/or materials with our publication.

## Abbreviations

ICIs: immune check point inhibitors
CTLA-4: cytotoxic T lymphocyte-associated protein 4
PD-1: programmed cell death protein 1
PD-L1: programmed cell death protein ligand 1
MSI: microsatellite instability
MMR: mismatch repair
HLA-I: human leukocyte antigen class I
OS: overall survival
TMB: tumor mutational burden
CR: complete response
PR: partial response
SD: stable disease
RECIST: response evaluation criteria in solid tumors
MDACC: M. D. Anderson Cancer Center
CM 012: CheckMate 012

