## Supplementary figures and images for "Association of HLA class I homozygosity with unfavorable clinical outcomes in patients with non-small cell lung cancer treated with PD-1/PD-L1 inhibitor as first-line therapy"

### Supplemental Figure A.1

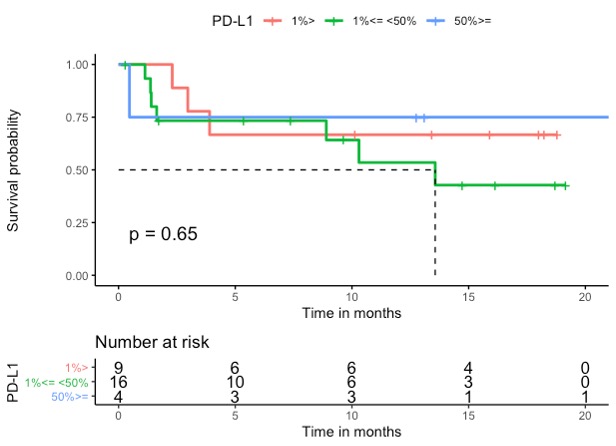

### Supplemental Figure A.2

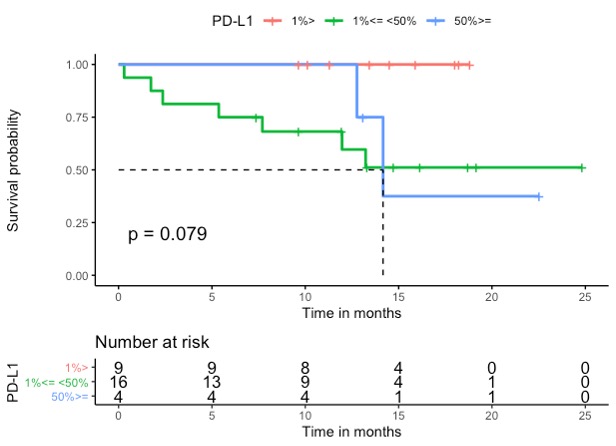

### Supplemental Figure B.1

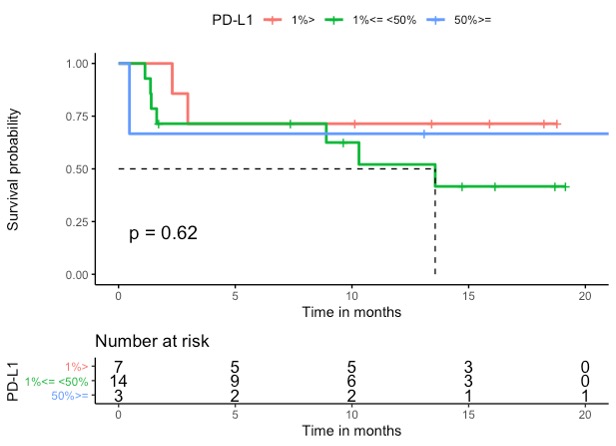

### Supplemental Figure B.2

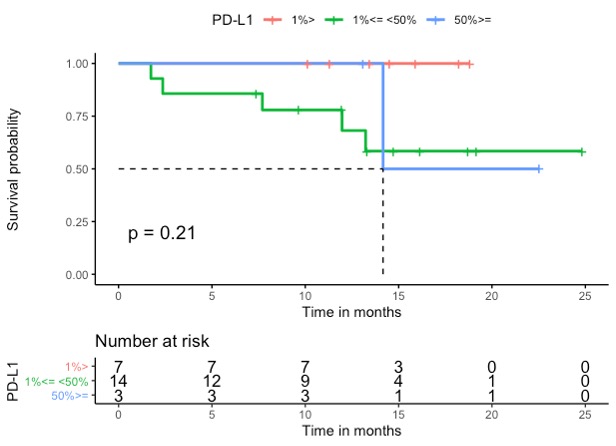

### Supplemental Figure C.1

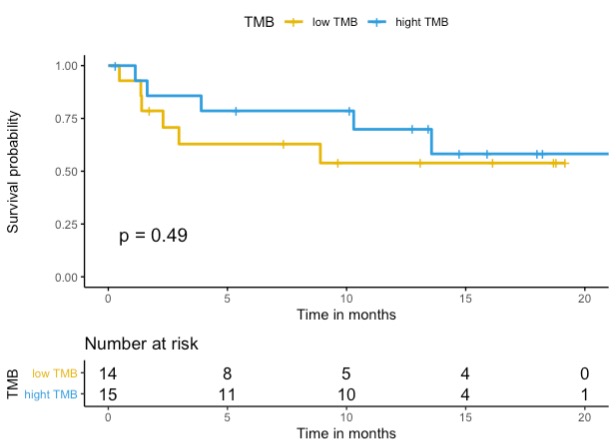

### Supplemental Figure C.2

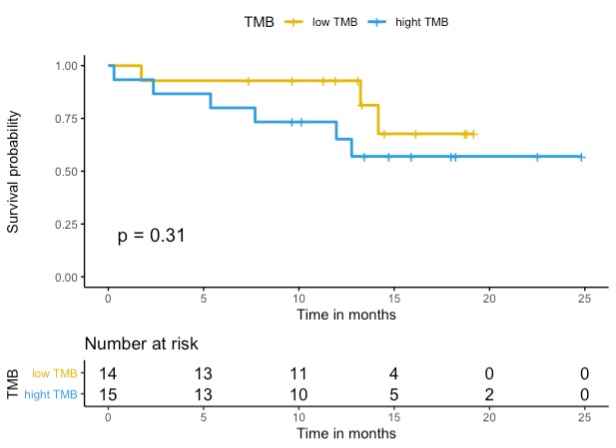

### Supplemental Figure D.1

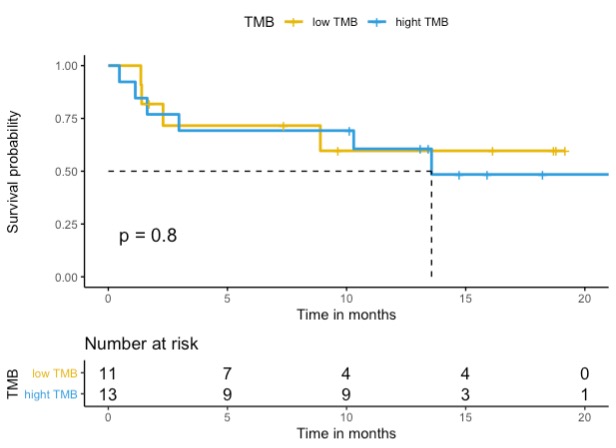

### Supplemental Figure D.2

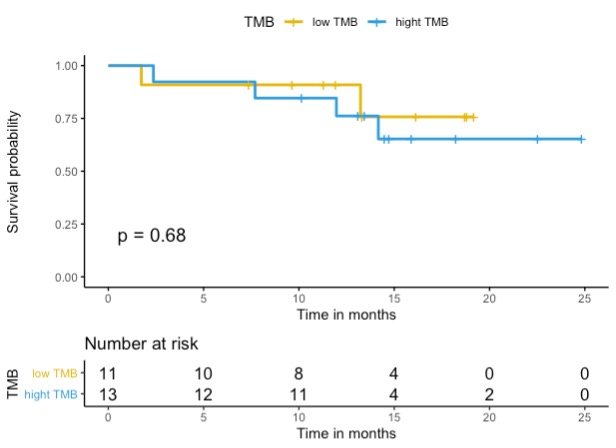

### Supplemental Figure E.1

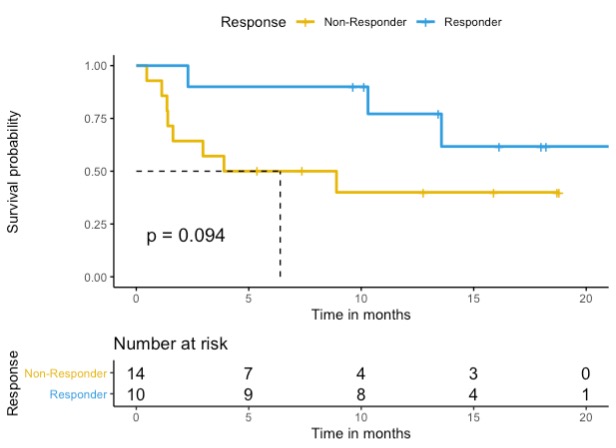

### Supplemental Figure E.2

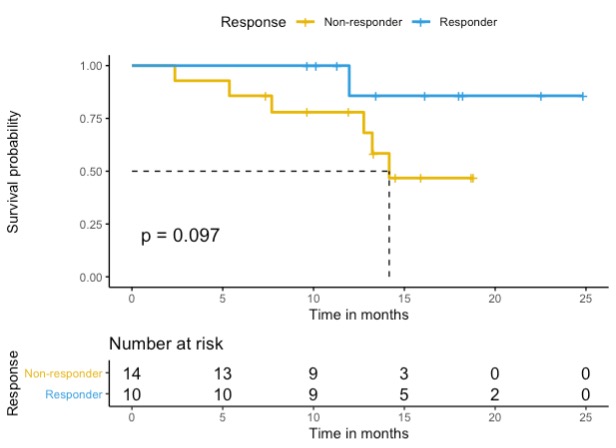
