## Supplemental Table A for "Association of HLA class I homozygosity with unfavorable clinical outcomes in patients with non-small cell lung cancer treated with PD-1/PD-L1 inhibitor as first-line therapy"

**Supplemental Table A. Characteristics of combination chemoimmunotherapy patient groups (N=24) with HLA-Heterozygous and HLA-Homozygous**

|  | HLA-Heterozygous  (N=20) | HLA-Homozygous  (N=4) | | P |
| --- | --- | --- | --- | --- |
| ***General characteristic*** |  |  | |  |
| Age, years | 67.0 ± 10.6 | 76.5 ± 6.2 | | 0.10 |
| Male, No. (%) | 10 (50.0%) | 2 (50.0%) | | 1.00 |
| BMI, kg/m^2^ | 25.0 ± 5.3 | 23.1 ± 1.5 | | 0.48 |
| Performance status |  |  | | 0.69 |
| - 0, No. (%) | 5 (25.0%) | 2 (50.0%) | |  |
| - 1-4, No. (%) | 15 (75.0%) | 2 (50.0%) | |  |
| Smoking, pack-year (%) | 37.2 ± 38.2 | 31.2 ± 46.1 | | 0.79 |
| COPD, No. (%) | 2 (10.0%) | 1 (25.0%) | | 1.00 |
| Lung cancer type |  |  | | 0.65 |
| - Non-squamous cell carcinoma, No. (%) | 15 (75.0%) | 4 (100.0%) | |  |
| - Squamous cell carcinoma, No. (%) | 5 (25.0%) | 0 (0.0%) | |  |
| Lung cancer stage |  |  | | 1.00 |
| - Stage 1-3, No. (%) | 5 (25.0%) | 1 (25.0%) | |  |
| - Stage 4, No. (%) | 15 (75.0%) | 3 (75.0%) | |  |
| Best overall response |  |  | | 0.34 |
| - Non applicable, No. (%)  - Non-CR/Non-PD, No. (%) | 4 (20.0%)  2 (10.0%) | 0 (0.0%)  0 (0.0%) | |  |
| - PD, No. (%) | 2 (10.0%) | 2 (50.0%) | |  |
| - PR, No. (%) | 5 (25.0%) | 1 (25.0%) | |  |
| - SD, No. (%) | 7 (35.0%) | 1 (25.0%) | |  |
| ***Laboratory findings*** |  | |  |  |
| Blood WBC count, x 10^9^/L | 9.2 ± 3.8 | 7.4 ± 2.7 | | 0.38 |
| Blood Hemoglobin, g/dL | 12.2 ± 2.0 | 12.9 ± 0.8 | | 0.50 |
| Blood Platelet count, x 10^9^/L | 352.4 ± 139.2 | 311.5 ± 39.0 | | 0.57 |
| Blood Neutrophil count, % | 72.9 ± 14.5 | 70.1 ± 13.6 | | 0.73 |
| Blood Lymphocyte count, % | 16.2 ± 11.3 | 18.5 ± 9.4 | | 0.70 |
| **Supplemental Table A. continued** |  |  | |  |
|  | HLA-Heterozygous  (N=20) | HLA-Homozygous  (N=4) | | P |
| ***Oncology testing panel findings*** |  |  | |  |
| PD-L1 (IHC) |  |  | | 0.50 |
| - <1%, No. (%) | 5 (25.0%) | 2 (50.0%) | |  |
| - 1%<= < 50%, No. (%) | 12 (60.0%) | 2 (50.0%) | |  |
| - 50%<=, No. (%) | 3 (15.0%) | 0 (0.0%) | |  |
| Tumor Mutation Burden, m/MB | 5.3 ± 3.4 | 2.3 ± 1.8 | | 0.11 |
| Neoantigen, No. (%) | 5.8 ± 5.5 | 1.8 ± 1.3 | | 0.01 |
| Frameshift mutations |  |  | | 0.25 |
| - 0, No. (%) | 8 (40.0%) | 1 (25.0%) | |  |
| - 1, No. (%) | 5 (25.0%) | 3 (75.0%) | |  |
| - 2, No. (%) | 5 (25.0%) | 0 (0.0%) | |  |
| - 3, No. (%) | 2 (10.0%) | 0 (0.0%) | |  |
| Abbreviations: BMI, body mass index; COPD, chronic obstructive pulmonary disease; PD, progressive of disease; PR, partial response; SD, stable disease; WBC, white blood cell; PD-L1, programmed death-ligand1; IHC, immunohistochemistry; | | | | |
