## Supplemental Table B for "Association of HLA class I homozygosity with unfavorable clinical outcomes in patients with non-small cell lung cancer treated with PD-1/PD-L1 inhibitor as first-line therapy"

**Supplemental Table B. HLA Class I genotype**

| ID | HLA-A diplotype | HLA-A supertype | HLA-B diplotype | HLA-B supertype | HLA-C diplotype |
| --- | --- | --- | --- | --- | --- |
| 1 | A*0201/A*2402 | A02/A24 | B*4601/B*4002 | B62/B44 | C*0102/C1502 |
| 2 | A*3601/A*6801 | A01/A03 | B*4403/B*1801 | B44/B44 | C*0401/C*0701 |
| 3 | A*3101/A*0201 | A03/A02 | B*4402/B*4001 | B44/B44 | C*0501/C*0304 |
| 4 | A*0101/A*0201 | A01/A02 | B*1501/B*0801 | B62/B08 | C*0304/C*0304 |
| 5 | A*2402/A*2601 | A24/A01 | B*2705/B*2705 | B27/B58 | C*0102/C*0701 |
| 6 | A*2417/A*1101 | A24/A03 | B*1502/B*1502 | B62/B62 | C*0801/C*0801 |
| 7 | A*3002/A*0202 | A01/A02 | B*1510/B*4102 | B27/B44 | C*1701/C*0304 |
| 8 | A*3402/A*3303 | A03/A03 | B*3501/B*1516 | B07/B58 | C*1402/C*0602 |
| 9 | A*3201/A*0205 | A01/A02 | B*5801/B*2702 | B58/B27 | C*0202/C*0701 |
| 10 | A*0201/A*3201 | A02/A01 | B*1302/B*2705 | Unclassified/B27 | C*0102/C*0602 |
| 11 | A*0201/A*2402 | A02/A24 | B*4402/B*4402 | B44/B44 | C*0202/C*0704 |
| 12 | A*0201/A*0101 | A02/A01 | B*2705/B*4403 | B27/B44 | C*0303/C*0102 |
| 13 | A*0201/A*0301 | A02/A03 | B*3501/B*0702 | B07/B07 | C*0701/C*0401 |
| 14 | A*0205/A*0201 | A02/A02 | B*4402/B*5101 | B44/B07 | C*0501/C*1402 |
| 15 | A*1101/A*0201 | A03/A02 | B*3501/B*0702 | B07/B07 | C*0702/C*0401 |
| 16 | A*0201/A*1101 | A02/A03 | B*5501/B*1501 | B07/B62 | C*0303/C*0303 |
| 17 | A*0203/A*1101 | A02/A03 | B*4801/B*3802 | B27/Unclassified | C*0702/C*0801 |
| 18 | A*3002/A*0205 | A01/A02 | B*5703/B*1503 | B58/B27 | C*0210/C*0701 |
| 19 | A*0201/A*3402 | A02/A03 | B*5301/B*3501 | B07/B07 | C*0401/C*0401 |
| 20 | A*3001/A*0201 | A01 A03/A02 | B*4403/B*1302 | B44/Unclassified | C*0602/C*1601 |
| 21 | A*0201/A*2402 | A02/A24 | B*1801/B*5501 | B44/B07 | C*0303/C*0701 |
| 22 | A*0101/A*2301 | A01/A24 | B*4403/B*0801 | B44/B08 | C*0701/C*0401 |
| 23 | A*0101/A*2402 | A01/A24 | B*0702/B*0801 | B07/B08 | C*0702/C*0701 |
| 24 | A*0101/A*2601 | A01/A01 | B*3501/B*0801 | B07/B08 | C*0701/C*0442 |
| 25 | A*2902/A*0201 | A01 A24/A01 | B*4402/B*4403 | B44/B44 | C*1601/C*0501 |
| 26 | A*2407/A*1101 | Unclassified/A03 | B*1502/B*3505 | B62/B07 | C*0702/C*0401 |
| 27 | A*0201/A*3303 | A02/A03 | B*5801/B*1302 | B58/Unclassified | C*0302/C*0602 |
| 28 | A*6801/A*0101 | A03/A01 | B*1801/B*3503 | B44/B07 | C*0401/C*1203 |
| 29 | A*0201/A*0101 | A02/A01 | B*4403/B*5501 | B44/B07 | C*0303/C*0401 |
