## Supplemental Table C for "Association of HLA class I homozygosity with unfavorable clinical outcomes in patients with non-small cell lung cancer treated with PD-1/PD-L1 inhibitor as first-line therapy"

|  | Patient No. (frequency) |
| --- | --- |
| ***HLA-A supertype*** |  |
| A01 | 14 (48.2) |
| A02 | 19 (65.5) |
| A03 | 12 (41.4) |
| A24 | 7 (24.1) |
| A01 A03 | 1 (3.4) |
| A01 A24 | 1 (3.4) |
| Unclassified | 1 (3.4) |
| ***HLA-B supertype*** |  |
| B07 | 12 (41.4) |
| B08 | 4 (13.8) |
| B27 | 7 (24.1) |
| B44 | 13 (44.8) |
| B58 | 5 (17.2) |
| B62 | 5 (17.2) |
| Unclassified | 4 (13.8) |

**Supplemental Table C. Patient groups classification with HLA supertypes (N=29)**
