## Supplemental Table D for "Association of HLA class I homozygosity with unfavorable clinical outcomes in patients with non-small cell lung cancer treated with PD-1/PD-L1 inhibitor as first-line therapy"

**Supplemental Table D. Hazard ratios for PFS for HLA-Heterozygous group vs HLA-Homozygous group (95% CI)**

|  | HLA-Heterozygous  (N=25) | HLA-Homozygous  (N=4) | P |
| --- | --- | --- | --- |
| Unadjusted (crude) | 1(ref) | 1.4 (0.30-6.4) | 0.68 |
| *Covariates included in the Cox model to calculate adjusted HRs* | | | |
| Age | 1(ref) | 1.5 (0.30-7.3) | 0.63 |
| Sex | 1(ref) | 1.4 (0.29-6.3) | 0.69 |
| BMI | 1(ref) | 1.4 (0.31-6.8) | 0.64 |
| COPD | 1(ref) | 1.4 (0.29-6.8) | 0.67 |
| Lung cancer type | 1(ref) | 1.1 (0.24-5.3) | 0.89 |
| Lung cancer stage | 1(ref) | 1.1 (0.24-5.3) | 0.89 |
| Performance status | 1(ref) | 1.5 (0.32-7.2) | 0.59 |
| Smoking, pack-year | 1(ref) | 1.3 (0.28-6.1) | 0.73 |
| Tumor Mutation Burden | 1(ref) | 1.1 (0.21-5.6) | 0.92 |
| PD-L1 (IHC) | 1(ref) | 1.5 (0.30-7.1) | 0.64 |
| Number of Neoantigen | 1(ref) | 1.1 (0.23-5.3) | 0.90 |
| Number of Frameshift mutations | 1(ref) | 1.5 (0.31-7.1) | 0.63 |
| Abbreviations: BMI, body mass index; COPD, chronic obstructive pulmonary disease; PD-L1, programmed death-ligand1; IHC, immunohistochemistry; | | | |
